# A Pseudotime-Dependent TWAS Framework Identifies Disease Genes along Cell Developmental Paths

**DOI:** 10.1101/2025.10.28.25338929

**Authors:** Rui Cao, Chunlin Li, Erjia Cui, Logan Spector, Andrew Raduski, Nathan Anderson, Weihua Guan, Peter Gordon, Cindy Im, Tianzhong Yang

## Abstract

Transcriptome-wide association studies (TWAS) link genes to disease risk by integrating gene expression with genome-wide association study (GWAS) data, where the use of bulk-tissue expression data typically provides gene-disease association interpretations at tissue levels. Recently, the increasing availability of single-cell gene expression profiles provides an opportunity to to dissect these associations at finer cellular granularity, allowing identification of cell-level effects that are not discernible from bulk-level analyses. While existing methods leverage single-cell data and map associations into discrete cell types, they may miss the continuous nature of cellular processes and misidentify causal cell stages in which genes exert their effects. To capture these continuous dynamic changes in gene expression, we developed the pseudotime-dependent Transcriptome-wide association study (pt-TWAS), a novel TWAS framework that captures gene effects along cell developmental paths and reveals their associations at a finer cell-stage resolution. By modeling gene expression as a continuous function of pseudotime, pt-TWAS gains statistical advantages over methods analyzing discrete cell types or stages. Specifically, it boosts statistical power by borrowing expression quantitative trait loci (eQTL) information across cell stages and jointly testing the gene-disease associations. Furthermore, pt-TWAS constructs and visualizes simultaneous confidence bands for the gene effect curve to identify the causal cell stage for the disease. As a demonstration of our method, we applied pt-TWAS to a GWAS of B-cell acute lymphoblastic leukemia (ALL) leveraging single-cell data from OneK1K, where we successfully replicated known risk genes from previous analyses and pinpointed their relevant cell stages. An R package implementing pt-TWAS is available at https://github.com/RuiCao34/ptTWAS/.

## 1 Introduction

Genome-wide association studies (GWAS) have identified thousands of genetic loci associated with complex human diseases (Buniello et al., 2019). However, because a majority of risk variants reside in non-coding regions of the genome, interpreting their biological mechanisms remains a notable challenge (Maurano et al., 2012; Watanabe et al., 2019). Transcriptome-wide association studies (TWAS) facilitate the interpretation by shifting the unit of association from individual genetic variants to genes (Gusev et al., 2016; Gamazon et al., 2015). As a gene-based method, TWAS leverages genetic variants as instruments to predict gene expression levels and then tests for associations between the predicted expression and a disease of interest. This approach not only boosts statistical power by aggregating the effects of multiple variants but also aligns with an instrumental variable (IV) regression framework, providing an approach for inferring potential causal links. The widespread success of this approach has been driven by large-scale public bulk-tissue expression Quantitative Trait Locus (eQTL) resources, such as the Genotype-Tissue Expression (GTEx) project (The GTEx Consortium, 2020). These resources provide pre-computed expression imputation models, enabling researchers with GWAS summary statistics to perform gene-based association tests and infer the genes’ roles in diseases (Wainberg et al., 2019).

While powerful, these analyses typically rely on static snapshots of gene expression and are heavily dominated by the common cell types in bulk tissue, which overlooks the rare cell types and the dynamic nature of genetic regulation, evidenced by recent findings (Yazar et al., 2022). This is a considerable limitation, as many diseases arise from perturbations of dynamic cellular processes like differentiation, activation, or response to stimuli (Hanahan and Weinberg, 2011; Wynn and Ramalingam, 2012; Umans et al., 2021) over a variety of cell stages. A key example and motivation of our study is acute lymphoblastic leukemia (ALL), the most common childhood malignancy, which is characterized by the malignant transformation and differentiation arrest of either B- or T-cell progenitors (Mullighan, 2012; Pagliaro et al., 2024). Given that the B-cell subtype accounts for the majority of ALL cases, its well-characterized developmental pathway from hematopoietic stem cells to mature plasma cells presents a clear modeling opportunity, particularly for the B cell stages present in accessible samples like peripheral blood. Identifying how dynamic genetic effects change throughout these developmental processes is therefore critical for designing more effective therapies. While current chimeric antigen receptor therapies (CAR-T), for instance, successfully target broadly expressed antigens like CD19 (Frey, 2019), next-generation strategies could achieve greater precision by targeting specific maturational stages where the disease process originates.

To move beyond bulk-tissue analysis, the advent of single-cell RNA sequencing (scRNA-seq) provides transcriptomic data with cellular-level resolution. However, adapting this high-resolution data for TWAS has often led to the creation of “pseudobulk” samples, where cells are aggregated within discrete, annotated cell types (Cuomo et al., 2023; Song et al., 2024). This approach fails to capture continuous cellular dynamics and may not pinpoint the causal cell stage, as eQTLs are frequently shared across cell stages (Cuomo et al., 2025) and a significant association in one binned cell stage could be driven by stronger correlated effects in a more causally relevant one. Furthermore, the pseudobulk approach is often underpowered for rare cell types due to limited numbers of cells.

A more powerful and flexible method for modeling cellular dynamics from cross-sectional scRNA-seq data is through pseudotime inference (Trapnell et al., 2014; Street et al., 2018). By ordering cells based on transcriptional similarity, pseudotime algorithms can reconstruct continuous cellular trajectories, providing a proxy for true temporal progression without the need for often-impractical longitudinal sampling. This enables us to move beyond analyses using binned cells (e.g., pseudobulk) and to model gene expression as a continuous function of a cell’s developmental state. We developed a novel pseudotime-dependent TWAS framework that incorporates single-cell transcriptomic data and disease GWAS data to infer causal genes at a cell-stage resolution. Statistically, pt-TWAS is a two-stage functional regression framework (**Figure 1**). In the first stage, we use a function-on-scalar regression model to predict the single-cell gene expression trajectory using local genetic variants. This stage jointly models cells across their trajectory and allows borrowing of eQTL information from more abundant cell types, increasing gene expression imputation accuracy. In the second stage, we test for an association between this imputed functional gene expression trajectory and an outcome. Specifically, we are interested in (1) a global test on whether there is any gene effect and (2) a subsequent non-flat test on whether there is any pseudotime-varying effect respectively. To infer the causal cell stage for a disease, we construct simultaneous confidence bands for the gene effect curve. As demonstrated in both our simulation and real data results, our method identifies dynamic and cell-stage-specific gene effects that can be missed by bulk-tissue or pseudobulk approaches. Our work has the potential to identify the precise cellular developmental window where a gene confers risk, which can guide the development of interventions targeting specific cell stages to prevent or mitigate disease gene effects.

**Figure 1:**
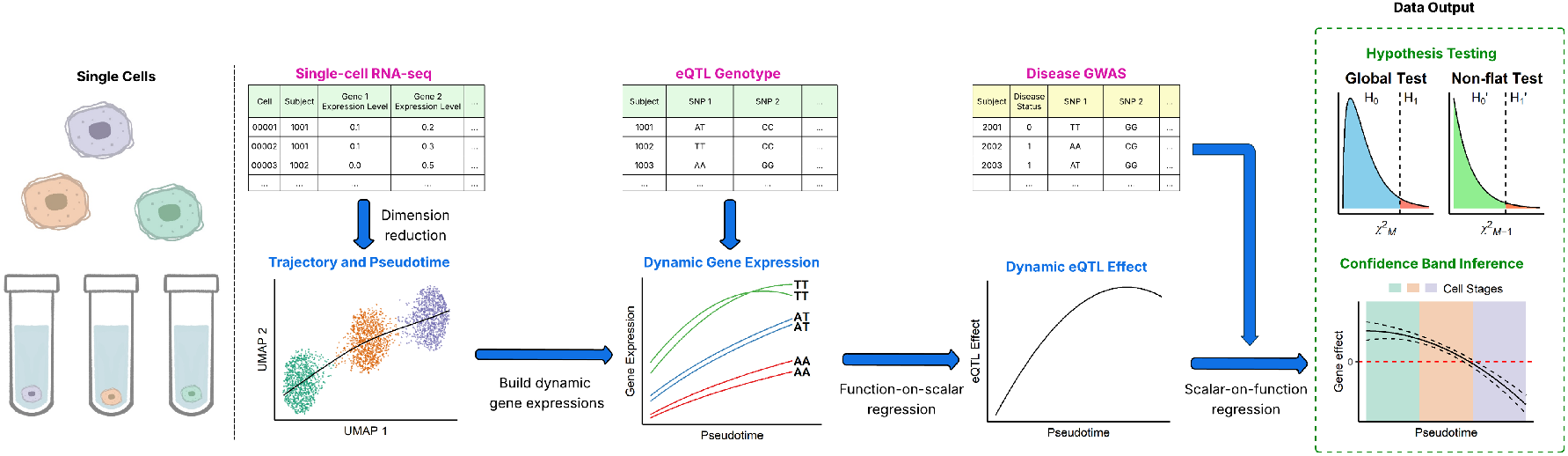
Overview of pt-TWAS framework. Starting from single-cell gene expression profiles and eQTL genotypes, gene expression imputation models were built through a function-on-scalar regression. Next, in an independent GWAS study, a scalar-on-function regression is carried out by regressing the GWAS outcome onto the genotype-predicted gene expression curve. Statistical tests for two null hypotheses (global and non-flat) were performed to assess the gene-outcome association. The estimate of the gene effect curve (solid black line in the bottom right figure) as well as its simultaneous confidence band (dashed black line) is constructed to identify the cell stages of the gene-disease association. 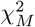 chi-squared distribution with degree of freedom *M* ; *M*, number of basis splines in the functional regressions.

## 2 Methods

### 2.1 Model Overview

Bulk-tissue TWAS usually consists of two stages: building an imputation model for gene expression within a tissue, i.e., averaged across cell types/stages, and testing its association with the disease of interest. In our framework, we develop a trajectory-based single-cell TWAS (pt-TWAS), where gene expression is modeled not as static but as a function of cell stages, quantified as pseudotime *t* inferred from the overall gene expression profiles of single cells. In Stage 1, we perform a function-on-scalar regression (Equation 5), fitting a linear mixed model with a group-lasso penalty to account for subject-level correlation and shrink the regression coefficients of SNPs with weak or no effect to zero, which is consistent with the empirical evidence that only a small number of cis-SNPs regulate gene expression in bulk tissue (Gamazon et al., 2015). In particular, the regression is fitted stepwise with respect to the degree of splines to control for smoothness and to prevent overfitting. In Stage 2, we perform a scalar-on-function regression (Equation 11) for the outcome-gene association. Multiple hypothesis tests can be conducted to draw inferences of interest. These include a global null hypothesis (no gene expression effect across all cell stages) and a secondary null hypothesis (no cell-stage-dependent effect). To infer the association strength and direction over cell stages, we construct simultaneous confidence bands for the gene effect curve. Similar to bulk-tissue TWAS, pt-TWAS can be applied to GWAS individual level data or summary statistics when a trained gene expression model and an external genotype reference panel are available (Methods regarding to the summary statistics in Supplementary Information Section S2). We compare our method to existing relevant TWAS methods and summarize it conceptually in **Table 3** with details in Section 4.

### 2.2 Stage 1 - Gene Expression Imputation Model

#### 2.2.1 Function-on-Scalar Regression

Suppose a single-cell data set consists of *S* subjects and *N*_*s*_ cells from subject *s*, with the total cell number 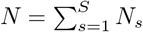. We treat the expression of a single gene of interest as a continuous variable (log-transformed read counts) (Hou et al., 2023), denoted by *x*_*sc*_ for the subject *s* and cell *c*, where *s* = 1, …, *S* and *c* = 1, …, *N*_*s*_. Let ***g***_*s*_ = (*g*_*s*1_, …, *g*_*sP*_ )^*T*^ be the genotype vector of *P* SNPs from subject *s* that are local to the gene of interest, ***u***_*s*_ = (*u*_*s*1_, …, *u*_*uQ*_)^*T*^ as *Q* subject-level covariates (e.g., sex, age, etc.), and *t ∈* [0, 1] as the inferred pseudotime along a single trajectory, representing a cell’s progression along a developmental trajectory, which we assume to be known. For subject *s* and cell *c*, we have

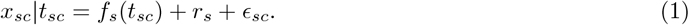

Assume the random intercepts 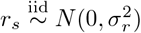 capture the within-subject correlation, the residuals 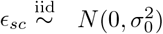, and the gene expression dynamics *f*_*s*_(*t*_*sc*_) can be modeled as a linear combination of *M* pre-specified basis splines ***b***(*t*_*sc*_) = (*b*_1_(*t*_*sc*_), …, *b*_*M*_ (*t*_*sc*_))^*T*^ with subject-specific weights ***w***_*s*_ = (*w*_*s*1_, …, *w*_*sM*_ )^*T*^

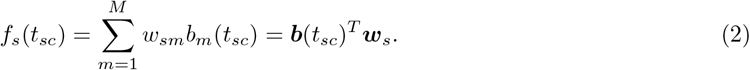

The weights *w*_*sm*_ are modeled as a linear combination of a baseline, genetic effects, and covariate effects.

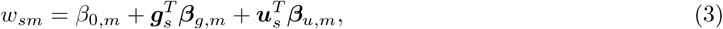

where *β*_0,*m*_ is the intercept, and ***β***_*g,m*_ = (*β*_*g,m*1_, …, *β*_*g,mP*_ )^*T*^ and ***β***_*u,m*_ = (*β*_*u,m*1_, …, *β*_*u,mQ*_)^*T*^ are the genotype and covariate effect coefficients, for the spline *m*. Rewrite ***w***_*s*_ in matrix form:

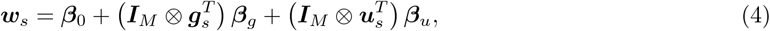

where *⊗* is the Kronecker product, and ***I***_*M*_ is an *M × M* identity matrix, ***β***_0_ = (*β*_0,1_, …, *β*_0,*M*_ )^*T*^, 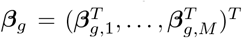, and 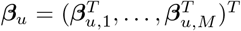. By simple algebra, the pseudotime-dependent Model 1 can be further rewritten as

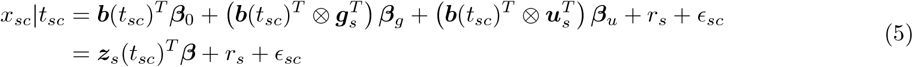

by defining 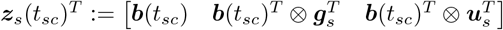 and 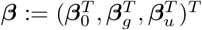.

#### 2.2.2 Parametric Estimation of *β*

In single-cell data, the gene expression in cells of subject *s*,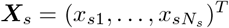, is observed at time points 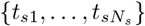. Stacking all *S* subjects, the full model can then be formulated as

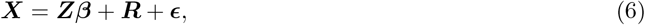

where 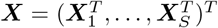, and ***Z, ϵ***, and ***R*** are the stacked forms of ***z***_*s*_(*t*_*sc*_), *r*_*s*_, and *ϵ*_*sc*_ respectively.

To reduce computational burden and alleviate the non-normality of *x*_*sc*_, which is characterized by excessive zeros in single-cell gene expression counts, we aggregate cells with similar pseudotime within the same subjects. This approach is in the spirit of single-cell aggregation methods (Baran et al., 2019). Within each subject, we cluster cells by Jenks natural breaks optimization by their pseudotime *t*, which minimizes the within-cluster variance and maximizes the between-cluster variance. We empirically set the number of clusters, *L*, to 10. This value prevents a heavy computational burden and lessens the zero-inflation issues, while also avoiding the loss of resolution that results from too few clusters. Following clustering, both the pseudotime and gene expression for each cell cluster are calculated as the mean values of the cells belonging to that cluster. We use the superscript *a* to denote cells after aggregation:

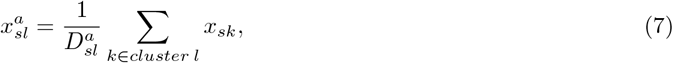

where 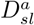 is the size of cluster *l* from subject *s*. Let 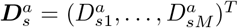 and 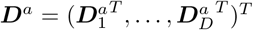.

Following the definition of ***X, Z, R***, and ***ϵ***, we have their aggregated form ***X***^*a*^, ***Z***^*a*^, ***R***^*a*^, and ***ϵ***^*a*^:

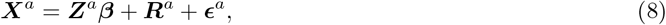

where 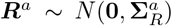 and 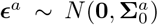. The variance of an aggregated observation is 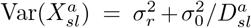. The covariance between two different aggregates for the same subject is 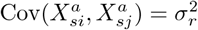 for *i ≠ j*. Therefore, the full variance-covariance matrix of the aggregated data, **Σ**^*a*^, is block-diagonal, with each variance block for each subject. The *s*^*th*^ block has diagonal elements 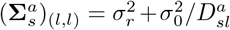 and off-diagonal elements 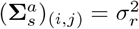. To estimate ***β*** in Equation 8, we fit the model with a group-lasso penalty to induce SNP-wise sparsity. We minimize the loss function

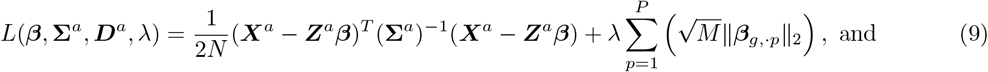

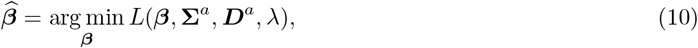

where ***β***_*g,·p*_ = (*β*_*g*,1*p*_, …, *β*_*g,Mp*_)^*T*^ is the regression coefficients for SNP *p* and *M* splines, and *λ* is the penalty parameter tuned by cross validation. Since the variance components 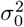 and 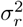 are usually unknown, we employ a two-step procedure to estimate ***β*** and **Σ**^*a*^ (**Algorithm 1**). First, we obtain an initial estimate of the coefficients, 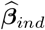, by fitting the group-lasso model assuming independent observations (i.e., setting 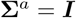). We then estimate the variance components 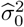 and 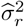 by applying Restricted Maximum Likelihood (REML) to the residuals from this initial fit 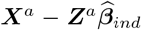. Second, we construct the estimated covariance matrix 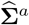 using the estimated variance components from REML. Its *s*^*th*^ block has diagonal elements 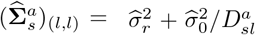 and off-diagonal elements 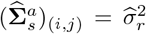. The final estimate 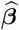 is then obtained by solving the group-lasso problem with correlated residuals, which is equivalent to decorrelating the outcome and predictors using 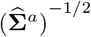 and fitting a standard group-lasso model. The REML algorithm and group-lasso regression are implemented using the R packages `lme4` (Bates et al., 2015) and `gglasso` (Yang and Zou, 2013) respectively.

#### 2.2.3 Choice of Spline Basis *b*(*t*)

Using single-cell gene expression data, we empirically find that orthonormal basis spline functions ***b***(*t*) up to the third order of *t* adequately model the gene expression curves. The candidate basis splines are derived through a Gram-Schmidt orthogonalization onto splines *{*1, *t, t*^2^, *t*^3^*}*. However, overfitting on the basis splines may lead to bias in the Stage 2 regression, analogous to the weak instrument (IV) problem documented in the two-stage least squares or Mendelian Randomization (MR) literature (Andrews et al., 2019; Shi et al., 2024). To ensure sufficient strength for IVs, analogous to multi-exposure IV regression (Sanderson and Windmeijer, 2016; Sanderson et al., 2019), we propose a forward stepwise procedure to control the number of fitted splines, where we examine the additional prediction power contributed by each basis spline conditional on the lower-order splines.

As shown in the **Algorithm 1**, we first regress the target gene expression onto the flat spline only *b*_1_(*t*) and get its cross-validated 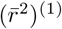 between predicted and observed outcomes. Next, we repeat the algorithm by including a higher-order spline *b*_2_(*t*) and get the cross-validated 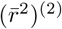. We keep the higher-order spline *b*_2_(*t*) in the prediction model only if the additional variance explained 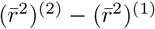 is larger than a pre-specified threshold. In lots of practices, *η* = 0.01 (Gamazon et al., 2015; He et al., 2022; Bhattacharya et al., 2021). The procedure above is repeated stepwise until an additional higher-order spline no longer results in an improvement in cross-validated 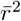.

Notably, our framework includes the standard pseudobulk imputation model as a special case. A pseudobulk analysis aggregates all cells from a given subject to derive a single, average expression value, thus disregarding the underlying cellular trajectory. This scenario arises within our framework if the stepwise selection procedure terminates after fitting the flat basis spline, that is, higher-order splines fail to provide a significant improvement in gene expression prediction. When only the flat basis spline is retained, the weak IV problem reduces to the standard case commonly studied in the literature (Andrews et al., 2019; Shi et al., 2024).

##### Algorithm 1: 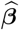 estimation in Stage 1.

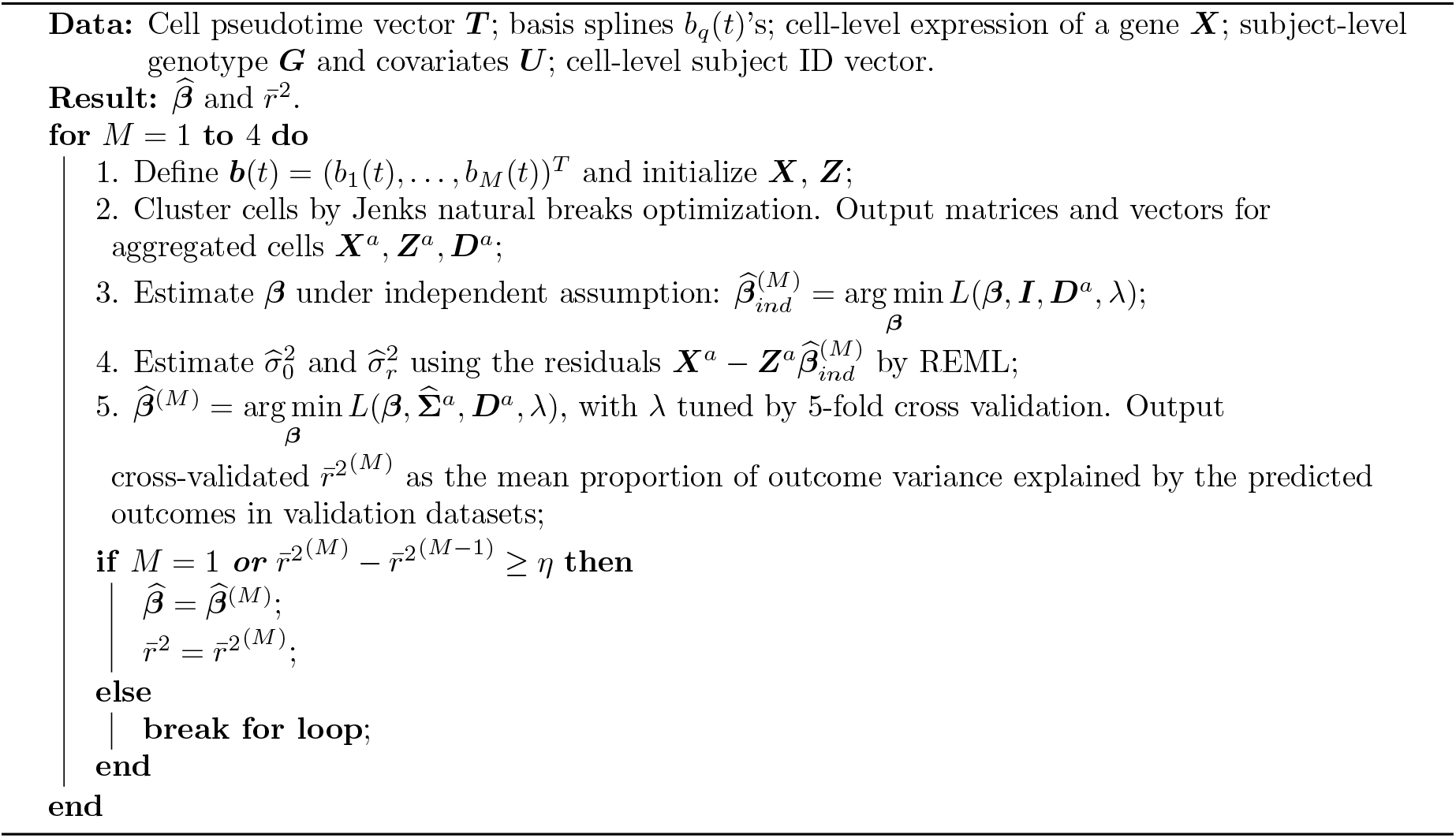

### 2.3 Stage 2 - Association Test with GWAS

#### 2.3.1 Scalar-on-Function Regression

Now we have the regression coefficient estimates 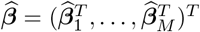. Suppose a GWAS dataset consists of *Ñ* subjects, with subjects that do not overlap with those in the Stage 1 single-cell dataset. The genotype matrix 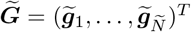 can be defined as a *Ñ × P* matrix with its (*s, p*)^*th*^ entry as the genotype of SNP *p* from subject *s*. Here we focus on a binary outcome ***Y*** = (*y*_1_, …, *y* _*Ñ*_)^*T*^ ; however, the method can be conveniently generalized to other types of outcomes. To test its association with the imputed gene expression, we model the total impact of gene expression on disease risk as an integral over the cell stages. Thus, we have the scalar-on-function regression model

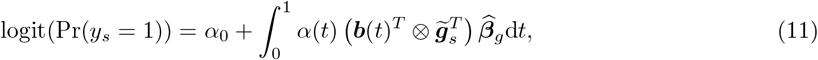

where *α*_0_ is the intercept effect and *α*(*t*) is the gene expression effect over pseudotime *t*. Similarly, we can decompose *α*(*t*) using several basis splines 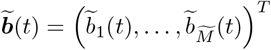:

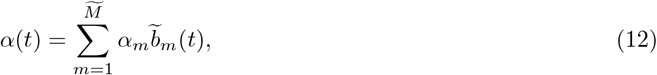

leading to

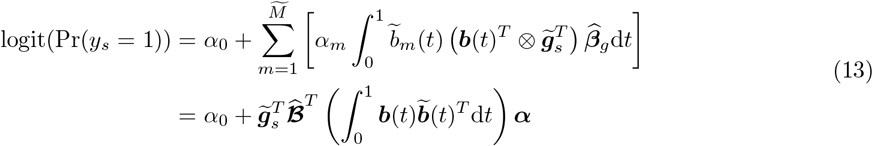

where ***α*** = (*α*_1_, …, *α*_*M*_ )^*T*^ is the regression coefficients and 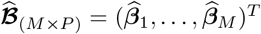 is the stacked form of 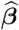. Practically, we use the same orthonormal splines as in Stage 1, based on the assumption that gene effect along the cell lineage is smooth. Under this case, the integral 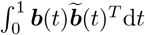 becomes an identity matrix, and the model takes the form of a standard logistic regression.

#### 2.3.2 Hypothesis Testing

As ***α*** is the spline coefficients for modeling *α*(*t*), we are interested in testing two null hypotheses: the global test and the non-flat test. For the global test, the null hypothesis *H*_0_ and its alternative *H*_1_ are defined as

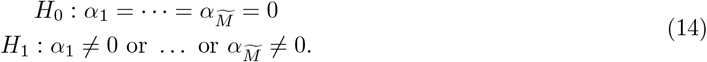

Rejecting *H*_0_ can be interpreted as a significant association between gene expression and the disease. For non-flat test, the null hypothesis 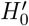 and its alternative 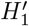 are

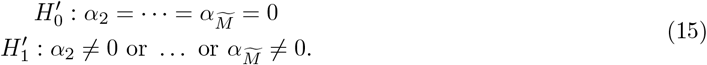

Note 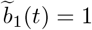 is the flat spline, that is, a pseudotime-invariant effect on the outcome. Rejecting 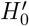 therefore indicates a significant pseudotime-varying association between gene expression and disease. The non-flat test may be of interest as it provides evidence on the time-varying gene effects that cannot be captured by existing pseudobulk methods. Both tests are based on Wald statistics, which asymptotically follow 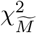 and 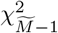 under their respective null hypotheses. To control the family-wise error rate of simultaneously conducting two tests, we use a hierarchical testing procedure: the global null hypothesis *H*_0_ is first tested at significance level *θ*, and the secondary non-flat hypothesis 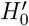 is tested at the same level only upon rejection of *H*_0_. This guarantees the overall type I error rate does not exceed *θ*.

Besides the global test and non-flat test, testing each single component of ***α*** may be relevant when 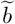 is a step function, as it is equivalent to testing the conditional cell-stage specific gene effect. A one degree of freedom Wald test can be applied here.

#### 2.3.3 Pointwise and Simultaneous Confidence Band Inference

If the global null hypothesis *H*_0_ is rejected, we are also interested in the inference about 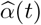, that is, at which specific cell stages the gene expression is associated with the disease. Under confidence level 1 *− θ*, the pointwise confidence band *h*_*p*_(*t*) can be straightforwardly derived from the regression coefficients 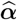

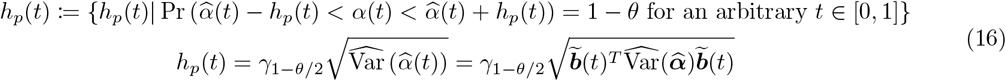

where *γ*_1*−θ/*2_ is the 1 *− θ/*2 quantile of a standard normal distribution, and 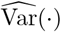 is the estimated variance-covariance matrix.

However, this approach focuses on the confidence intervals of individual points and does not take into account the multiple comparisons over *t ∈* [0, 1]. Thus we also construct a simultaneous confidence band *h*_*s*_(*t*) in our study, defined as

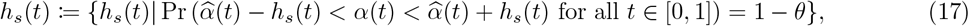

We construct the simultaneous confidence band using a parametric bootstrap method (Crainiceanu et al., 2024; Hall and Horowitz, 2013), which empirically controls the simultaneous coverage probability by testing it over a fine grid of points on the interval *t ∈* [0, 1], as detailed in **Algorithm 2**. We infer a significant gene-disease association exists in any pseudotime interval where the simultaneous confidence band does not contain zero.

##### Algorithm 2: Parametric bootstrap for simultaneous confidence bands.

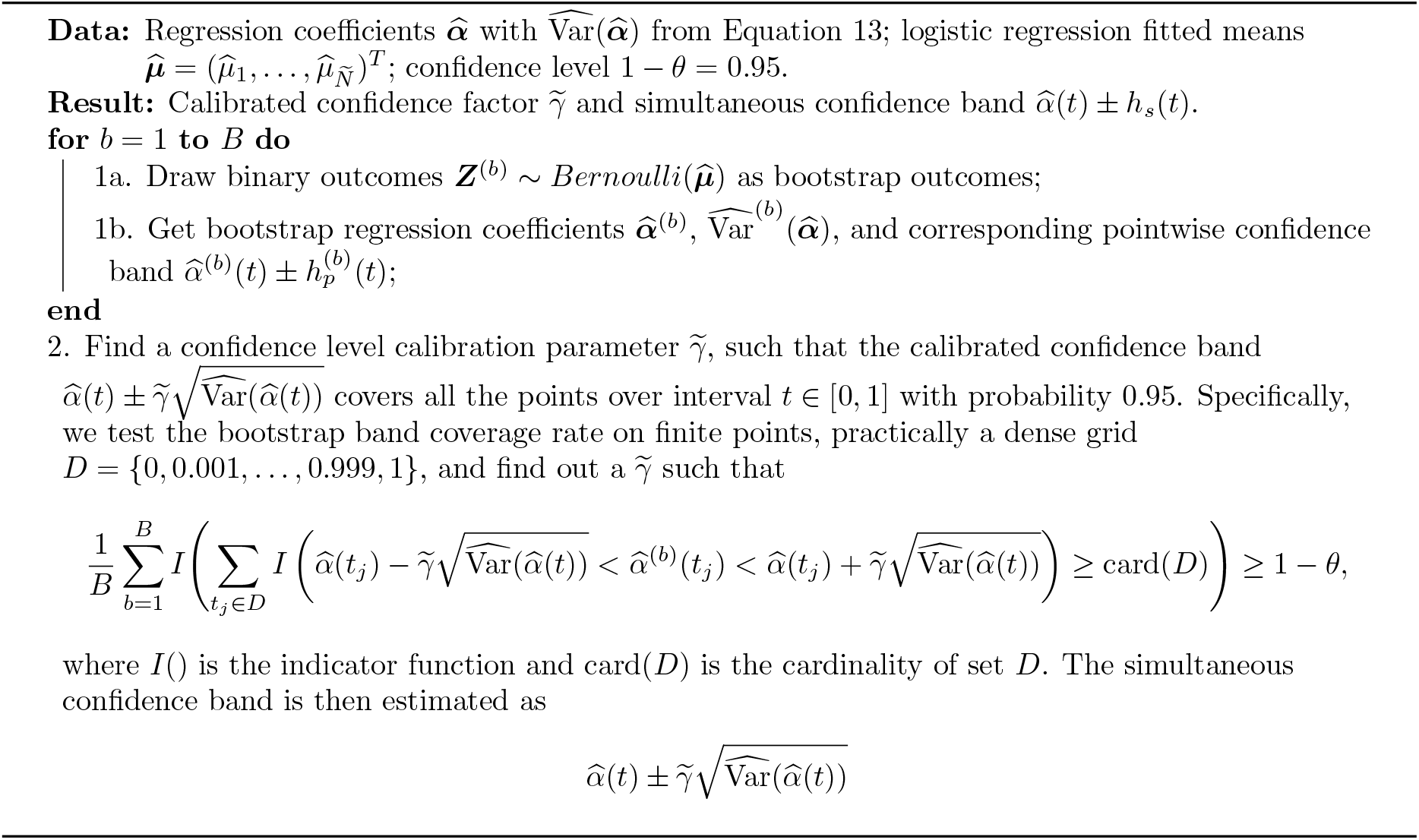

To facilitate interpretation, we scale the association effect curve 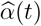 by multiplying by a factor of the average gene expression standard deviation over *t*. The standardized effect 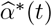 is calculated as

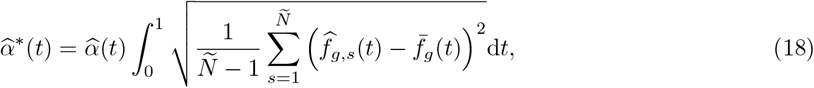

where 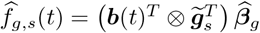 is the genetic component of expression for subject *s*, and 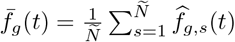 is its average among *Ñ* subjects. After the normalization, 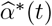 is interpreted as the effect curve under a one-standard-deviation change in the overall predicted gene expression. Since 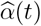 and 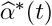 are always proportional over *t*, the simultaneous confidence band of 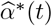 can be straightforwardly derived from the simultaneous confidence band of 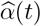.

#### 2.3.4 Choice of Spline Basis 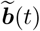

In Section 2.3.1, our Stage 2 regression model assumes the same smooth basis splines as in Stage 1, that is,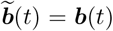. This provides flexibility to approximate dynamic associations to continuous smooth curves. Herein, we discuss the impact of potential model misspecification (i.e., choice of wrong spline basis) in Stage 2 regression, with respect to the dimension of Stage 2 spline basis 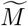.

- 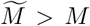: When the Stage 2 basis dimension is greater than the Stage 1 dimension, the regression model is not identifiable because the regression matrix 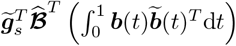 is not of full column rank (Supplementary Information). In other words, when the variation of the gene expression curve is captured by *M* splines, its effect curve *α*(*t*) cannot be modeled by a higher number of coefficients. This is analogous to a conventional IV regression where the number of exposures cannot exceed the number of IVs.
- 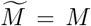: A natural choice in Stage 2 is to use the same orthonormal splines as those in Stage 1, reflecting the assumption that the gene effect *α*(*t*) is continuous and smooth. Alternatively, one could use a different spline basis 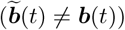 of the same dimension. For example, the use of step functions as basis splines in Stage 2 assumes the gene effect is constant within discrete cell stages. We show that in this case, the P-value of the global test remains invariant to the choice of basis (proof in Supplementary Information Section S1 and empirical evidence in Section 2.4), although the estimated confidence bands depend on it. For the purpose of identifying causal cell stages, we recommend sensitivity analyses with alternative bases (e.g., step functions). Consistency of the inferred stages across spline bases would indicate greater robustness.
- 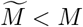: It is also feasible to use a lower-dimensional spline basis in Stage 2 regression. For instance, gene expression can be modeled as dynamic across cell stages, while its effect on the outcome is assumed to be pseudotime-invariant 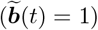. This approach is distinct from a standard pseudobulk analysis, which is a special case of our framework where both the gene expression and its effect are modeled as constant 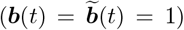. More generally, using fewer basis functions in Stage 2 can increase statistical power when higher-order effects are weak or absent, since tests incorporating more splines lose efficiency due to the additional degrees of freedom.

### 2.4 Simulation Studies for pt-TWAS

We performed extensive simulations to evaluate the type I error and power for the global and non-flat tests both under correct model specification and model misspecification. Furthermore, we benchmarked our method with pseudobulk TWAS across the whole cell stages and specific cell-stages of interest. In addition to identifying genes of interest, we also examined the performance of identifying causal stages using the simultaneous confidence band beyond simple visualization.

To assess the type I error of pt-TWAS, we simulated two scenarios under the global null hypothesis of no association (*α*(*t*) = 0). In the literature, single cell gene expression is commonly modeled in two ways, either as a continuous variable (e.g., after log transformation) (Lin et al., 2022; Hou et al., 2023) or as counts with or without explicit modeling of excessive zeros (Hafemeister and Satija, 2019; Hou et al., 2023; Xi and Li, 2023). Scenario 1 assumed data conformed to our model’s assumptions, with the gene expression outcome *x*_*sc*_ simulated as a continuous variable from Model 5. Scenario 2 evaluated the robustness under model misspecification of the gene imputation model (Stage 1) by generating continuous values as in Scenario 1, then exponentiating and flooring them into integers, thereby introducing excess zeros. Note that in Scenario 2, we evaluated the type I error control under both violations of the linearity and residual normality assumptions.

We also compared the empirical power of our method with pseudobulk TWAS, where the gene expression level was the mean of cell-level expressions per individual. Specifically, the pseudobulk approach applied a Lasso penalty term for SNP-wise sparsity in the gene imputation model and regressed the outcome onto the scalar gene expression. We considered *α*(*t*) as a linear (Scenario 3) or quadratic function (Scenario 4) of *t*, testing our method under various gene effects over cell stages. In Scenario 5, *α*(*t*) was simulated as a step function, mimicking a biological process where the gene is causal for a disease within only one specific cell stage. In this scenario, we also compared the statistical power with cell-stage-specific pseudobulk TWAS.

In all simulation scenarios, the gene had *P* = 50 eQTLs and *Q* = 5 covariates in the single cell dataset (Stage 1). Five of the 50 eQTLs and all of the five covariates had non-zero effects on gene expression. SNPs were generated from an independent binomial distribution with frequency 0.4, and covariates were drawn independently from standard normal distributions *N* (0, 1). The residuals came from independent standard normal distributions 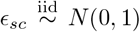. In stage 2, we simulated *Ñ* = 2, 000 GWAS subjects, with genotypes following the same distributions as in Stage 1. The binary outcomes were simulated from the logistic regression model, where the intercept term was set to achieve an approximate 1:1 case-control ratio. Type I error and statistical power of hypothesis tests were estimated as the empirical probability of rejecting the global or non-flat null hypotheses across 1,000 repetitions. The power of confidence band inference is defined as the probability that the simultaneous confidence band does not cover 0 at some pseudotime point within a given interval. The details of each scenario are described below.

- **Scenario 1**. Each iteration consisted of *S* = 500 subjects and *N*_*s*_ = 50 cells per subject, resulting in a total cell sample size *N* = 25, 000. The cell pseudotime was independently drawn from the uniform distribution *U* (0, 1). Subject-level intercept *r*_*s*_ = 0, *x*_*sc*_ was simulated as a continuous outcome *x*_*sc*_|*t*_*sc*_ = ***z***_*s*_(*t*_*sc*_)^*T*^ ***β*** + *r*_*s*_ + *ϵ*_*sc*_, baseline gene expression curve 0.5*b*_1_(*t*) + *b*_2_(*t*) *− b*_3_(*t*), flat-spline eQTL effects 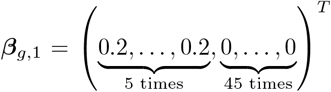, linear-spline eQTL effects 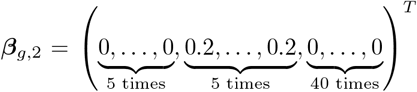, quadratic-spline eQTL effects 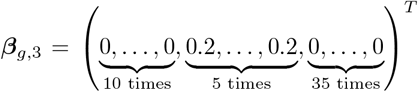, flat-spline, linear-spline, and quadratic covariate effects 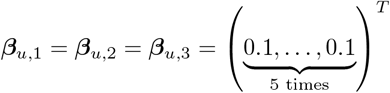, gene effect *α*(*t*) = 0 when *t ∈* [0, 1].
- **Scenario 2**. Each iteration consisted of *S* = 500 subjects and *N*_*s*_ = 50 cells per subject, resulting in a total cell sample size *N* = 25, 000. The cell pseudotime was independently drawn from the uniform distribution *U* (0, 1). Subject-level intercept *r*_*s*_ = 0.1, *x*_*sc*_ was simulated as a count outcome *x*_*sc*_|*t*_*sc*_ = ⌊exp(***z***_*s*_(*t*_*sc*_)^*T*^ ***β*** + *r*_*s*_ + *ϵ*_*sc*_)⌋, where ⌊·⌋ was the floor function, baseline gene expression curve *−*0.5*b*_1_(*t*) + *b*_2_(*t*), flat-spline eQTL effects 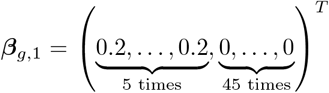, linear-spline eQTL effects 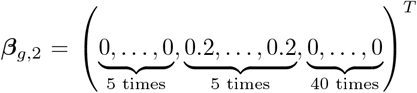, flat-spline and linear-spline covariate effects 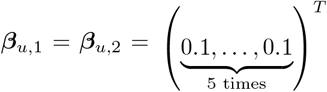, gene effect *α*(*t*) = 0 when *t ∈* [0, 1].
- **Scenario 3**. Each iteration consisted of *S* = 500 subjects and *N*_*s*_ = 50 cells per subject, resulting in a total cell sample size *N* = 25, 000. The cell pseudotime was independently drawn from the uniform distribution *U* (0, 1). Subject-level intercept *r*_*s*_ = 0, *x*_*sc*_ was simulated as a continuous outcome *x*_*sc*_|*t*_*sc*_ = ***z***_*s*_(*t*_*sc*_)^*T*^ ***β*** + *r*_*s*_ + *ϵ*_*sc*_, baseline gene expression curve 0.5*b*_1_(*t*) + *b*_2_(*t*), flat-spline eQTL effects 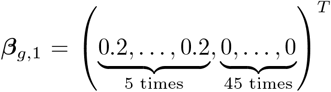, linear-spline eQTL effects 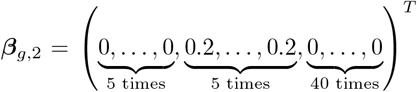, flat-spline and linear-spline covariate effect 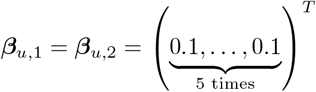, gene effect *α*(*t*) = 0.2*b*_1_(*t*)+ 0.2*b*_2_(*t*) when *t ∈* [0, 1]. In addition to using the flat and linear spline bases, we also examined model misspecification in Stage 2 using step function bases, where 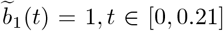 and 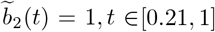, corresponding to the cell stages with negative or positive gene effects.
- **Scenario 4**. Each iteration consisted of *S* = 500 subjects and *N*_*s*_ = 50 cells per subject, resulting in a total cell sample size *N* = 25, 000. The cell pseudotime was independently drawn from the uniform distribution *U* (0, 1). Subject-level intercept *r*_*s*_ = 0, *x*_*sc*_ was simulated as a continuous outcome *x*_*sc*_|*t*_*sc*_ = ***z***_*s*_(*t*_*sc*_)^*T*^ ***β*** + *r*_*s*_ + *ϵ*_*sc*_, baseline gene expression curve 0.5*b*_1_(*t*) + *b*_2_(*t*) *− b*_3_(*t*), flat-spline eQTL effects 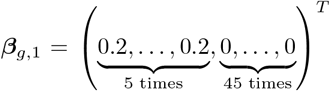, linear-spline eQTL effects 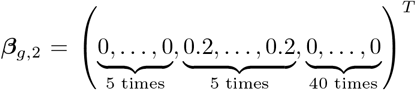, quadratic-spline eQTL effects 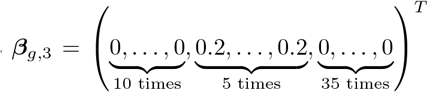, flat-spline, linear-spline, and quadratic covariate effect 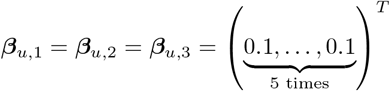, gene effect *α*(*t*) = 0.1*b*_2_(*t*) + 0.2*b*_3_(*t*) when *t ∈* [0, 1].
- **Scenario 5**. Each iteration consisted of *S* = 100 subjects and *N*_*s*_ = 110 cells per subject, where for each individual 10 cells were uniformly distributed in an earlier stage *t ∈* [0, 0.8] and 100 cells uniformly distributed in a later stage *t ∈* [0.8, 1], resulting in a total cell sample size *N* = 11, 000. Subject-level intercept *r*_*s*_ = 0, *x*_*sc*_ was simulated as a continuous outcome *x*_*sc*_|*t*_*sc*_ = ***z***_*s*_(*t*_*sc*_)^*T*^ ***β*** +*r*_*s*_ +*ϵ*_*sc*_, baseline gene expression curve 0.5*b*_1_(*t*) + *b*_2_(*t*), flat-spline eQTL effect 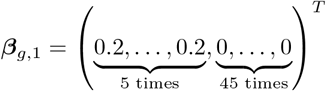, linear-spline eQTL effects 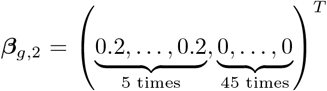, quadratic-spline eQTL effects ***β***_*g*,3_ = **0**, flat-spline and linear-spline covariate effects were **0**, gene effect

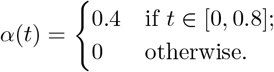

### 2.5 Real Data Analysis

#### 2.5.1 Single-cell Gene Imputation Models

The single-cell gene imputation models were derived from the OneK1K study (Yazar et al., 2022), to date the largest public single-cell peripheral blood mononuclear cell (PBMC) dataset. It consists of 1,102 individuals of Northern European ancestry and single-cell RNA-seq data from 1,267,768 PBMCs of 32,738 genes. After genotype calling and imputation, 4,749,893 SNPs from 23 chromosomes were genotyped and imputed for the subjects. The single-cell RNA-seq data were cleaned according to Yazar et al. (2022). Since we focuses on B-cell ALL, we annotated these PBMCs using R package `Azimuth` (Hao et al., 2021) and identified 112,098 naïve and memory B cells combined. For the B cells, we inferred a single trajectory from the naïve B-cell stage to the memory B-cell stage, where the pseudotime was the cell rank along the trajectory after normalization to [0, 1]. The trajectories and pseudotime were inferred using `slingshot` (Street et al., 2018). Details for the OneK1K genomic data cleaning and processing can be found in Supplementary Information Sections S3-S5.

#### 2.5.2 GWAS: B-cell Acute Lymphocytic Leukemia

ALL is a childhood cancer of the blood and bone marrow, characterized by overproduction and proliferation of abnormal immature lymphocytes. B-cell ALL (B-ALL), specifically, is the predominant ALL subtype arising when B cell proliferation becomes uncontrolled. Previous GWAS and TWAS (Chokkalingam et al., 2013; Xu et al., 2013; Migliorini et al., 2013; Ellinghaus et al., 2012; Yang et al., 2022) have identified several dozens of genetic loci related to B-ALL, but it remains unknown at which stage of B-cell development these genes initiate or influence ALL. In this study, we investigated 17 candidate genes previously reported associated with B-ALL, and applied pt-TWAS to identify the cell stages of these associations. For method comparison, we also applied a pseudobulk approach, which aggregated the individual-level gene expressions in B cells. The B-ALL individual-level GWAS data comes from the SMILES study (Geris et al., 2023), consisting of 842 cases and 1,570 controls of European ancestry. The cases were children aged 0 to 14 years between 1987 and 2014 in Michigan with a B-ALL diagnosis, and the controls were matched to cases under a case-control ratio of 1:2. The main result of pt-TWAS will be presented under the orthonormal spline basis, i.e. continuous functions. As a sensitivity analysis, we analyzed the significant genes using a step spline basis, modeling abrupt changes in gene effects across developmental cell stages.

## 3 Results

### 3.1 Simulation Studies

We conducted simulation studies with five distinct scenarios to evaluate the performance of our method. The first two scenarios assessed the type I error rate under the null hypothesis of no association (*α*(*t*) = 0). Scenario 1 simulated gene expression as a continuous outcome directly from our proposed model (Equation 6), representing an ideal setting. Scenario 2 simulated sparse count-based expression, with an average zero proportion of 36.7%. In both scenarios, the empirical type I errors for the pt-TWAS global and non-flat tests, as well as for the pseudobulk TWAS, were well-controlled at the significance level 0.05 (**Figure 2A**). The results aligned with our expectation as the outcome ***Y*** was independent of the design matrix when the true effect *α*(*t*) was zero, as demonstrated in Model (11).

**Figure 2:**
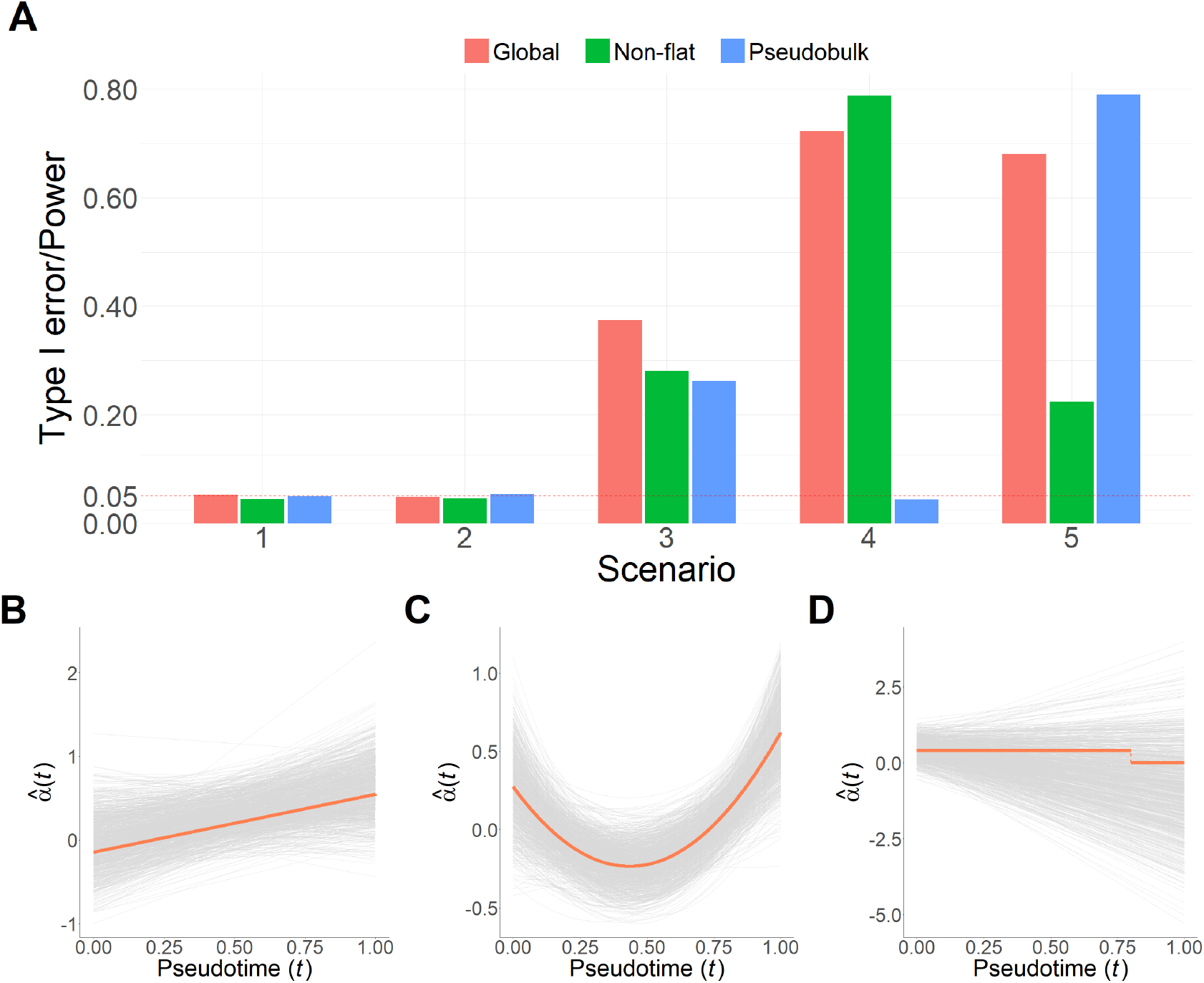
Simulation results for pt-TWAS and pseudobulk TWAS. A) Type I error (Scenarios 1 and 2) and statistical power (Scenarios 3, 4, and 5) for the pt-TWAS and pseudobulk TWAS; B,C,D) 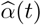 over simulation iterations (light gray lines) and true effect curve *α*(*t*) (solid orange lines) in scenarios 3, 4, and 5. The pseudobulk method aggregates all the cells within *t ∈* [0, 1].

In Scenarios 3 and 4, we evaluated the statistical power of pt-TWAS relative to the pseudobulk TWAS. In Scenario 3, both the underlying gene expression trajectory *f*_*s*_(*t*) and the true effect curve *α*(*t*) were linear functions of pseudotime. In Scenario 4, we simulated a more complex non-linear association where both functions were quadratic. In both settings, pt-TWAS achieved higher power than the pseudobulk approach (**Figure 2A**). The power advantage was particularly pronounced in Scenario 4, where the true effect was positive at the beginning and end of the trajectory but negative in the middle. This nonlinear pattern of *α*(*t*) resulted in the effects largely cancelled out when aggregated in the pseudobulk TWAS analysis, leading to a near-complete loss of power. Furthermore, our estimated effect curves, 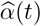, successfully recovered the true functional form of the association in both power scenarios (**Figure 2B,C**), which contributed to its higher power than pseudobulk TWAS.

In Scenario 5, the true effect *α*(*t*) was a discrete step function, where the gene had a constant effect over the earlier cell stage *t ∈* [0, 0.8] but no effect over the later cell stage *t ∈* [0.8, 1]. Specifically, the cell abundance between the two cell stages was imbalanced (ratio 1:10), with the earlier stage occurring more rarely. Using linear smooth splines 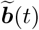 in Stage 2 regression, pt-TWAS approximated the true effect *α*(*t*) as linear curves (**Figure 2D**). Consistent with the proof in the Supplementary Information Section S1, we observed that the power of the Wald-statistic-based global test was not affected by the choice of splines 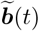, i.e., using linear basis vs the correct step function. While the pt-TWAS global test was less powerful than the standard pseudobulk test (0.680 vs 0.790), it outperformed a pseudobulk analysis restricted to the earlier stage (0.523), where only earlier-stage cells were used in the gene imputation model. Notably, the pt-TWAS global P-value is invariant to the range of cell stages in the Stage 2 (*t ∈* [0, 1] or [0, 0.8]) because it remains invariant with regression splines 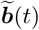. This demonstrates that pt-TWAS’s increased statistical power stems from a more accurate gene imputation model, built by leveraging a larger cell sample to estimate cross-stage eQTL effects. Additionally, the type I error for a later-stage-specific (*t ∈* [0.8, 1]) pseudobulk analysis was inflated (0.648), driven by the strong correlation in the imputed gene expression between the two cell stages. An analogous situation occurs in GWAS, where linkage disequilibrium can produce spurious signals at non-causal SNPs. Therefore, such marginal cell-stage-specific pseudobulk analyses may not be ideal for identifying causal cell stages.

We evaluated the performance of simultaneous confidence band inference in identifying causal cell stages in Scenarios 3, 4, and 5. As shown in **Table 1**, it had higher power to identify causal cdll stages with stronger effects across all scenarios. The analysis had higher power when using discrete step splines, which aggregate cell stages with the same causal direction, than when using smooth splines. In Scenario 5, the discrete step splines successfully controlled the type I error rate, in contrast to the marginal cell-stage-specific pseudobulk analysis, showcasing its superiority to identify causal cell stages over pseudobulk TWAS analysis.

**Table 1:** Simulated statistical power of confidence band inference using different splines.

| Simulation Scenario | Cell Stage ($t$ interval) | Gene Effect Direction | Strongest Gene Effect within an Interval | Statistical Power | |
| --- | --- | --- | --- | --- | --- |
|  |  |  |  | Smooth Orthonormal Splines | Discrete Step Splines |
| 3 | [0, 0.21] | - | -0.15 | 0.020 | 0.035 |
|  | [0.21, 1] | + | 0.55 | 0.336 | 0.420 |
| 4 | [0, 0.14] | + | 0.27 | 0.066 | 0.086 |
|  | [0.14, 0.73] | - | -0.23 | 0.207 | 0.251 |
|  | [0.73, 1] | + | 0.62 | 0.509 | 0.553 |
| 5 | [0, 0.8] | + | 0.4 | 0.339 | 0.356 |
|  | [0.8, 1] | /* | 0 | 0.005 | 0.055 |

\* No gene effect in this interval, and type I errors are shown in this row.

### 3.2 Real Data Analysis

Using the OneK1K single-cell data, we inferred the differentiation path of B cells. The B cell analysis revealed a single, linear trajectory spanning from naïve to memory B cells (**Figure 3**). Of the 17 candidate genes evaluated, our analysis using pt-TWAS identified eight that passed the heritability cutoff (pt-TWAS *h*^2^ *>* 0.01). This resulted in a Bonferroni-corrected significance threshold of 6.25 *×* 10^*−*3^. Applying this cutoff, we identified two genes, *IKZF1* and *COMMD3*, with expression patterns significantly associated with B-ALL.

**Figure 3:**
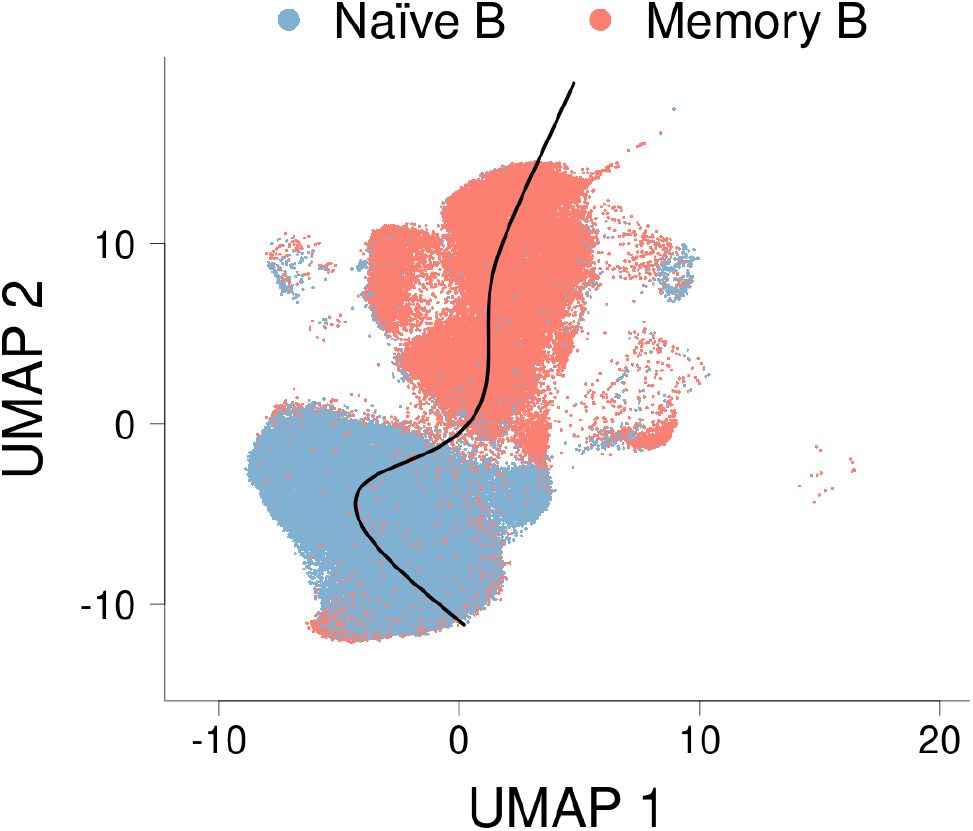
Trajectories inferred from B cells in OneK1K. The black solid curve indicates the cell developmental path from the naïve stage (bottom) to the memory stage (top).

The most significant gene identified was *IKZF1*, which showed a strong global association with B-ALL (global P-value = 1.71 *×* 10^*−*3^). While its dynamic effect across B cell development was not statistically significant after multiple testing correction (non-flat P-value = 1.42 *×* 10^*−*2^), the estimated effect strength decreased from the naïve to the memory B-cell stage (**Figure 4A**). Critically, the confidence band for its effect revealed a significant negative association specifically at the naïve B cell stage. *IKZF1* is a well-established B-ALL risk gene (Marke et al., 2018; Mullighan et al., 2009; Paolino et al., 2024), but to our knowledge, this is the first evidence pinpointing its association to naïve peripheral blood B cells, demonstrating the enhanced resolution of our method.

**Figure 4:**
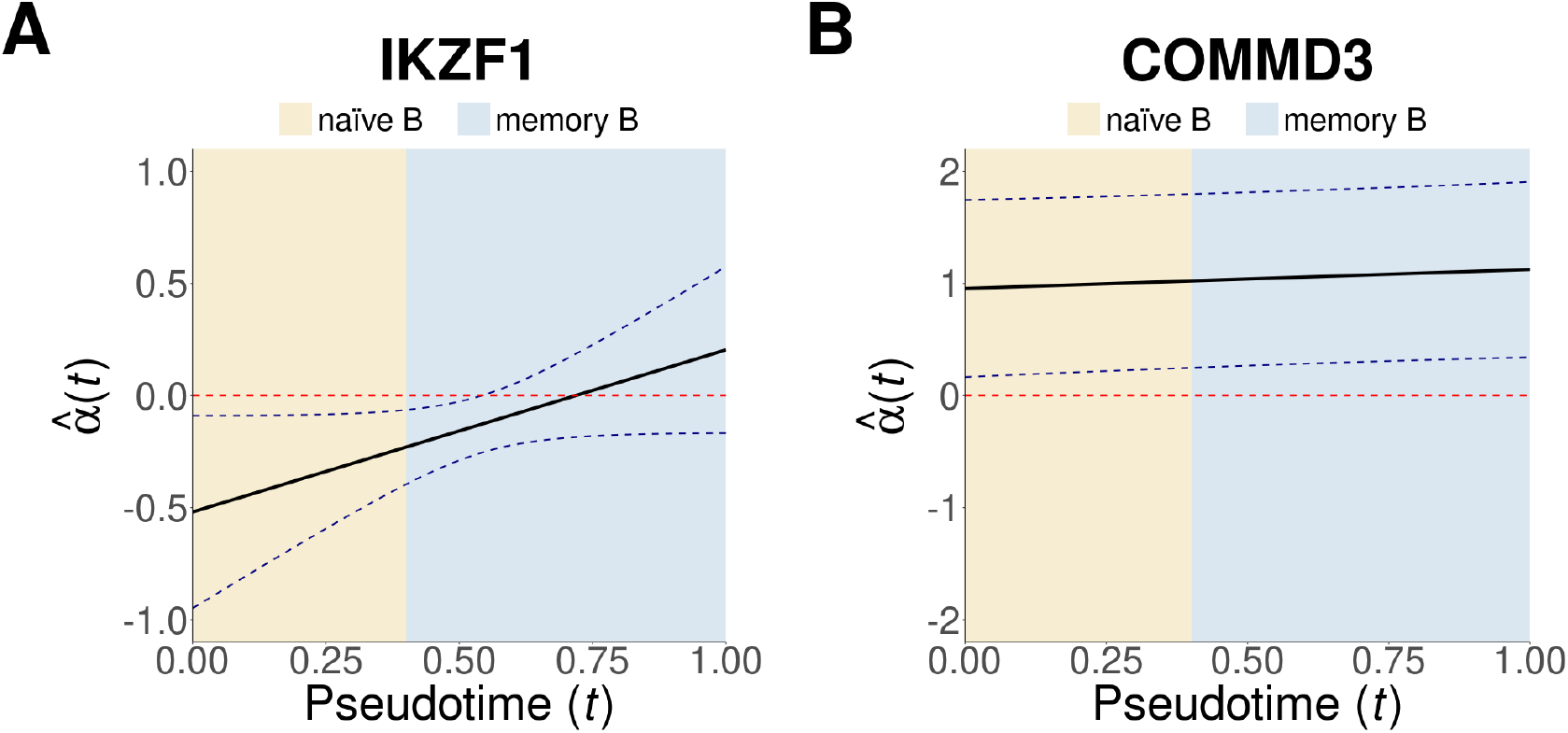
*IKZF1* (left) and *COMMD3* (right) confidence bands on B-ALL. Solid black line,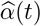; dashed blue line, simultaneous confidence band; dashed red line, horizontal line at 0; beige color, naïve B cell stage; light blue color, memory B cell stage.

The second significant gene, *COMMD3*, also demonstrated a global association with B-ALL (global P-value = 5.42 *×* 10^*−*3^). In contrast to *IKZF1*, its effect was relatively constant across B-cell developmental stages (non-flat P-value = 1.94 *×* 10^*−*1^). The confidence band indicated a consistent positive association throughout both naïve and memory stages (**Figure 4B**). This finding aligns with previous reports of *COMMD3* overexpression in ALL patients (Mulaw et al., 2012) and its relevance in B-cell lymphomas (Ferreira et al., 2008).

As a sensitivity analysis, we estimated the simultaneous confidence bands using step functions for ***b***(*t*) (**Figure S1**). The inferred causal cell stages were consistent with those found using smooth orthonormal splines. This result demonstrated the robustness of our research findings in the B cell lineage.

We also performed a standard pseudobulk analysis using the same B cells as in pt-TWAS. In this analysis, only one gene, *PIP4K2A*, passed the heritability cutoff. The poorly imputed pseudobulk models were also noted in a recent TWAS publication using OneK1K single-cell data (Zhou et al., 2025). The higher number of imputable genes observed in pt-TWAS was likely because our method effectively captures dynamic eQTL effects that are averaged out in pseudobulk analyses. Although the *PIP4K2A* association with B-ALL did not reach statistical significance (P-value = 0.072), the positive direction of its effect (estimate = 0.07) is consistent with previous reports (Zhang et al., 2019; Liao et al., 2016). The non-significant P-value for *PIP4K2A* is likely attributable to low statistical power, which might be resolved with a larger B-ALL GWAS.

## 4 Discussion

In this work, we introduced pt-TWAS, a novel framework that integrates the principles of TWAS with single-cell data by modeling gene expression as a continuous function of pseudotime. Our framework is implemented as a two-stage functional regression. In the first stage, we build a predictive model for the gene expression trajectory using a parametric function-on-scalar mixed model, with a group-lasso penalty to enforce SNP-wise sparsity. In the second stage, we carry out a scalar-on-function regression between a disease outcome and the imputed expression trajectory. This allows us to formally test global and dynamic gene-disease associations, from which confidence bands can be constructed to infer the cell stages driving the association. Similar to existing TWAS methods, the data used in the two stages are not required to be collected on the same subjects. To further broaden the use of our framework, we show that the second-stage regression is also compatible with GWAS summary statistics (Supplementary Information SectionS2), which allows it to be readily applied to vast public datasets without requiring individual-level genotype data. The algorithm was implemented at https://github.com/RuiCao34/ptTWAS/. Herein, we highlight several key modeling considerations in developing the pt-TWAS framework. These include ensuring sufficient instrument strength for reliable gene expression imputation, appropriately specifying the functional basis for pseudotime-dependent effects, and accurately modeling single-cell expression measurements.

A key assumption for TWAS is the instrumental relevance, i.e., genetic variants/instruments explain enough variance in the gene expression imputation model. Analogous to the “weak instrument” problem in conventional TWAS, that is, weak instruments can bias causal estimates in the Stage 2 regression, pt-TWAS can also be biased from such weak instruments. Our framework addresses this challenge with a two-tiered selection strategy (detailed in Section 2.2.3). First, at the SNP level, the group-lasso penalty selects for genetic variants that have non-zero effects on the expression trajectory, effectively excluding weak or noisy SNPs from the model. Second, at the functional level, our forward stepwise procedure selects only those splines that contribute to the gene expression prediction. The stopping criterion for this procedure, a minimum increase in cross-validated R-squared, ensures the final imputed expression trajectory is adequately predicted by the genetic variants without overfitting.

Another important modeling decision in single-cell RNA-seq analysis is how to handle the gene expression measurements. Raw scRNA-seq count data typically exhibit high sparsity. Statistically, count-based distributions like the Negative Binomial or its zero-inflated variants provide the most accurate representation of this data structure (Pan et al., 2023; Lin et al., 2022). However, fitting these complex, non-linear models is computationally intensive, particularly within the penalized, mixed-effects framework required for our analysis. A common and more tractable alternative is to apply a log-transformation to the counts, which stabilizes variance and enables the use of the well-established linear models. Our decision to aggregate cells into small pseudotime-based clusters before log-transforming represents a pragmatic compromise: it reduces the severity of zero-inflation, making the Gaussian assumption more tenable, while preserving the high resolution of the trajectory and ensuring the model remains computationally efficient. Importantly, our simulations demonstrate that this choice is robust, as our framework successfully controls the type I error even when the underlying data are generated from a count distribution with excessive zeros.

Our framework offers distinct advantages over bulk-tissue and pseudobulk methods in both stages of analysis (Comparison summarized in **Table 3**). In the first stage, conventional bulk or pseudobulk TWAS imputes average gene expression and estimates an average gene effect across all cell stages. While cell-stage-specific pseudobulk analysis can test effects within discrete stages, it is prone to misidentifying causal gene stage due to expression correlation between them. In contrast, pt-TWAS models gene expression as a continuous trajectory. By leveraging the shared nature of eQTL effects across the cellular continuum, our model borrows information from abundant cell populations to improve expression imputation for rarer cell states, which are often poorly predicted in the pseudobulk analyses. In the second stage, pt-TWAS performs a joint test on the entire imputed trajectory, unlike pseudobulk methods that conduct a series of marginal tests on each stage. By analyzing the full expression lineage, our joint test has greater power to detect dynamic associations. This allows it to more accurately pinpoint the true causal cell stage and avoid the signal misidentification common in marginal analyses where expression levels are correlated across stages.

**Table 2:** pt-TWAS and pseudobulk analysis for genetically imputable B-ALL genes using smooth orthonormal splines.

| Gene | Chr | Gene Start<br>(bp) | Gene End<br>(bp) | Global<br>P-value | Non-flat<br>P-value | cis-SNPs | Non-zero<br>SNPs | # splines | $\hat{h}^2$<br>pt-TWAS | $\hat{h}^2$<br>pseudobulk |
| --- | --- | --- | --- | --- | --- | --- | --- | --- | --- | --- |
| *IKZF1* | 7 | 50,304,068 | 50,405,101 | $1.71 \times 10^{-3}$ | $1.42 \times 10^{-2}$ | 240 | 124 | 2 | 0.022 | 0.009 |
| *COMMD3* | 10 | 22,316,386 | 22,320,306 | $5.42 \times 10^{-3}$ | $1.94 \times 10^{-1}$ | 174 | 22 | 2 | 0.015 | 0.006 |
| *ARID5B* | 10 | 61,901,684 | 62,096,944 | $1.81 \times 10^{-1}$ | $7.69 \times 10^{-2}$ | 290 | 190 | 2 | 0.082 | 0.004 |
| *PIP4K2A* | 10 | 22,534,854 | 22,714,578 | $3.66 \times 10^{-1}$ | $6.67 \times 10^{-1}$ | 160 | 66 | 2 | 0.032 | 0.012 |
| *C14orf119* | 14 | 23,095,505 | 23,100,456 | $3.81 \times 10^{-1}$ | $2.50 \times 10^{-1}$ | 202 | 64 | 2 | 0.022 | 0.007 |
| *LHPP* | 10 | 124,461,823 | 124,617,888 | $5.55 \times 10^{-1}$ | $3.38 \times 10^{-1}$ | 384 | 116 | 2 | 0.034 | 0.006 |
| *INTS10* | 8 | 19,817,391 | 19,852,083 | $5.66 \times 10^{-1}$ | $7.03 \times 10^{-1}$ | 692 | 124 | 2 | 0.014 | 0.002 |
| *CDKN2A* | 9 | 21,967,752 | 21,995,301 | $6.30 \times 10^{-1}$ | $9.67 \times 10^{-1}$ | 455 | 43 | 2 | 0.052 | 0.001 |
| **Chr**, Chromosome; **bp**, base pair (GRCh38); **Non-zero SNPs**, SNPs with non-zero effects in the imputation model; **# splines**, number of basis spline functions; **$\hat{h}^2$ pt-TWAS**, cross-validated gene heritability fitted by pt-TWAS; **$\hat{h}^2$ pseudobulk**, cross-validated gene heritability fitted by pseudobulk TWAS. |  |  |  |  |  |  |  |  |  |  |

**Table 3:** Comparison of pseudobulk TWAS, cell-stage-specific pseudobulk TWAS, and pt-TWAS methods.

| Method | Estimand | Connection with pt-TWAS | Comments |
| --- | --- | --- | --- |
| Pseudobulk | Average gene effect $\alpha$ (scalar) | Equivalent to pt-TWAS when $\mathbf{b}(t) = \tilde{\mathbf{b}}(t) = 1$ for $t \in [0, 1]$ | • Identifies causal genes by averaging effects across all cell stages<br>• Cannot localize effects to specific cell stages<br>• Loses power when effects vary across stages |
| Cell-stage-specific pseudobulk | Marginal cell-stage gene effect $\alpha$ (scalar) | Equivalent to pt-TWAS when $\mathbf{b}(t) = \tilde{\mathbf{b}}(t) = 1$ for some $t$ interval $[c_1, c_2]$. | • Estimates the marginal cell-stage gene effect<br>• Prone to false causal genes due to cross-cell-stage gene expression correlation |
| pt-TWAS (smooth gene effect) | Gene effect coefficients $\alpha$ onto splines | Smooth splines $\mathbf{b}(t) = \tilde{\mathbf{b}}(t)$. | • Models both gene expression and its effect as smooth curves<br>• Infers causal cell stages using the confidence band |
| pt-TWAS (discrete gene effect) | Gene effect coefficients $\alpha$ onto cell stages | Smooth splines $\mathbf{b}(t)$ and discrete splines $\tilde{\mathbf{b}}(t)$. | • Models smooth gene expression and discrete (step-wise) gene effects<br>• More powerful if cell stages are correctly defined and at lower risk of identifying the incorrect cell stages than cell-stage-specific pseudobulk<br>• Useful for sensitivity analysis |

We also explored the impact of model misspecification, specifically the choice of spline functions in the Stage 2 regression. We proved that while the global P-value is invariant to the splines used in Stage 2, the confidence band inference for localizing causal cell stages depends on this choice. Furthermore, simulations demonstrated that using step functions can increase statistical power, provided that gene effects are unidirectional within pre-defined cell stages. In practice, however, identifying these cell stage intervals can be infeasible sometimes and can compromise the power and type I error for detecting the true causal stages. Given this dependency, applying pt-TWAS with alternative spline configurations serves as an important sensitivity analysis.

Several limitations of our analysis should be acknowledged. First, inferring causal genes through pt-TWAS inherits the assumptions of two-stage least squares frameworks such as TWAS and MR. Valid causal interpretation requires that the genetic instruments (1) are strongly associated with gene expression, (2) influence the outcome only through the target gene (no horizontal pleiotropy), and (3) are independent of unmeasured confounders. While our two-tiered instrument selection procedure helps address the first assumption and mitigates weak-instrument bias, the latter two are difficult to verify. In particular, horizontal pleiotropy remains a potential source of bias. Therefore, our findings should be interpreted as robust gene–trait associations rather than definitive causal effects. Additionally, the current single-cell eQTL sample sizes may limit the accuracy of gene expression imputation, though ongoing efforts to generate larger and more diverse datasets (Cuomo et al., 2025) are expected to improve both power and reliability.

Second, the inference of causal cell stages is subject to modeling choices. The confidence bands used to identify these stages depend on the spline basis used in Stage 2 of pt-TWAS. A gene’s true effect trajectory can be smooth or exhibit abrupt changes, and the optimal spline basis is difficult to determine *a priori*. Smooth splines may obscure sharp, stage-specific effects and reduce statistical power, whereas discrete step functions risk identifying false positives if stage boundaries are specified incorrectly. Therefore, while our primary analysis uses smooth orthonormal splines, we acknowledge that the identified causal windows could shift under different assumptions, underscoring the importance of conducting sensitivity analyses to ensure robust conclusions. Moreover, inferring causal cell stage is also dependent on the single cell dataset used in Stage 1. In our real analysis, we inferred causal cell stage for B-ALL using single-cell data from peripheral blood, whereas the primary site of leukemogenesis is the bone marrow; thus potentially limit our findings. Future studies using bone marrow–derived cells will provide a more direct characterization of disease-relevant developmental stages.

Looking forward, several avenues exist for extending this framework. Our work focuses on single trajectory, however, an extension of our model to handle complex, branching lineages may be of interest. Another important area for future research is addressing the invalid instrument issue and generalizing to robust MR methods. Extending MR to a setting with a “functional exposure” like an expression trajectory presents statistical challenges. Furthermore, our current framework assumes that the pseudotime ordering is known and accurate. In reality, pseudotime inference has its own sources of uncertainty, and future work could focus on developing methods to account for this uncertainty throughout the pt-TWAS pipeline. Despite these limitations, the generalizability of pt-TWAS is a major strength. Because pt-TWAS is fully compatible with GWAS summary statistics, it can be readily applied to hundreds of existing and future GWAS datasets for a wide range of human diseases and traits. To facilitate broader adoption, we have developed an open-source R package implementing pt-TWAS, which supports both individual-level and summary-level data. This will enable the broader scientific community to dissect the cellular dynamics of genetic risk and translate GWAS findings into precise and mechanistic insights.

## Data Availability

All data produced in the present work are contained in the manuscript

## Acknowledgment

R. Cao gratefully acknowledges support from the Doctoral Dissertation Fellowship by the University of Minnesota and thanks Ms. Chang Li for her skillful assistance in drawing Figure 1. T. Yang is supported by the St. Baldrick Career Award (1463960) and NIH grant R01AG074858. C. Li is supported in part by NSF grant DMS-2515789. The authors acknowledge the Minnesota Supercomputing Institute (MSI) at the University of Minnesota for providing resources that contributed to the research results reported within this paper. URL: http://www.msi.umn.edu. The authors thank the OneK1K project for sharing the single-cell data.

## Data Availability

OneK1K single-cell gene expression and genotype data were downloaded from Gene Expression Omnibus (GSE196830).

## Supplementary Information

### S1 Candidate Spline Basis in Stage 1 and 2 regression

In Stage 1 function-on-scalar regression, the candidate orthonormal basis splines are derived through Gram-Schmidt orthogonalization of the polynomial basis *{*1, *t, t*^2^, *t*^3^*}*

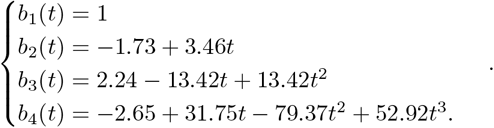

In stage 2 scalar-on-function regression, we found the number of basis splines in stage 2 cannot exceed that in Stage 1. Intuitively speaking, the higher-order effect *α*(*t*) is non-identifiable if the gene expression curve is modeled using a lower-order basis. This is analogous to the relationship between instrumental variables (IVs) and exposure in multi-exposure IV regression, where the number of exposures cannot exceed the number of IVs. As shown in Model 21, the rank of the design matrix 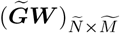 is limited by the rank of the matrix 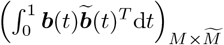. When 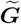 and 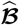 are full-rank, the rank of the design matrix is determined by the inner product of the basis functions, i.e. rank 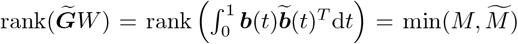. Thus the design matrix 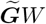 is not of full-rank if 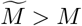.

To prove the global P-value is invariant with the choice of 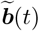, let **Γ** denote the integral matrix 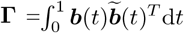 in the regression model 13. As long as **Γ** is a full-rank square matrix, the column space of the design matrix 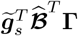 and the fitted means 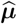 with its variance-covariance estimate 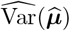 are invariant to **Γ**, that is, the choice of 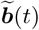. The regression coefficients are estimated as 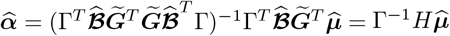, where 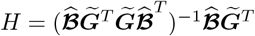. Simple algebra further shows the Wald statistic 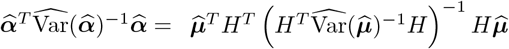, which is invariant with **Γ**. Consequently, the P-value of the global test is independent of this choice. A similar conclusion holds for the P-value of the non-flat test, which is the P-value of the remaining splines conditionally on *α*_1_ for the first spline *α*_1_(*t*) = 1. This is analogous to the property in generalized linear regressions that the joint significance of a set of covariates is invariant to a full-rank linear transformation of those covariates.

### S2 Extension to GWAS Summary Statistics

Similar to conventional TWAS, the method can work with GWAS summary statistics and an external reference panel without requiring the individual-level genotypes 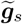. For the regression using summary statistics, we start with the linear regression model for continuous outcomes ***Y*** = (*y*_1_, …, *y*_*s*_)^*T*^ and residuals 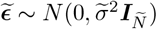 and then generalize to logistic regression:

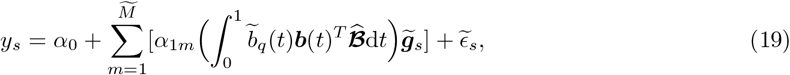

or in the matrix form:

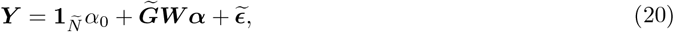

where **1**_*Ñ*_ is the one vector of length *Ñ*, 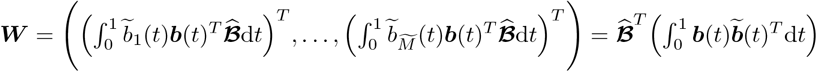 and 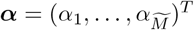. Since the intercept effect *α*_0_ is usually not of interest nor reported in GWAS summary statistics, we assume the outcome ***Y*** has been regressed out of the intercept and we have regression model

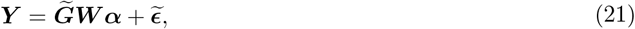

and its estimates:

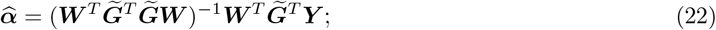

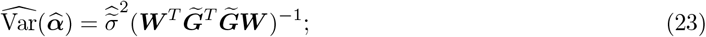

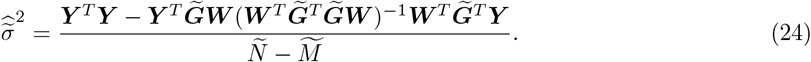

Suppose the GWAS summary statistics consist of regression coefficients estimates 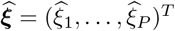 and standard errors 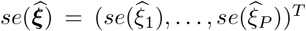. The terms 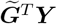 and ***Y*** ^*T*^ ***Y*** can be derived from GWAS summary data (Pattee and Pan, 2020):

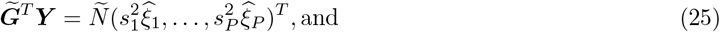

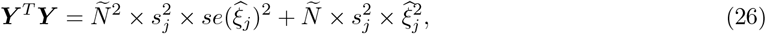

where 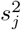 is the variance of SNP *j*. Both 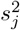 and 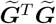 terms can be estimated from a reference panel. Practically, ***Y*** ^*T*^ ***Y*** can be estimated by the median over a given set of SNPs when ***Y*** is not standardized. The primary hypothesis can be then tested using the F-test on 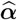, and the secondary null hypothesis by testing the sub-vector with *α*_1_ removed.

For a binary outcome, we can approximate the logistic regression coefficients and 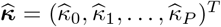 and standard errors and 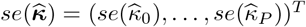 by

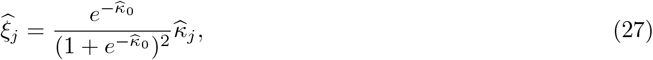

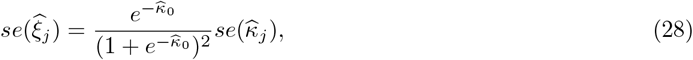

where *j* = 1, …, *P* and 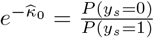 is the control-case ratio when assuming the 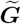 columns are centered at 0. The logistic regression coefficients can be thus converted to linear regression coefficients and handled by the steps mentioned above. After the regression, the coefficients are transformed back to the logistic scale by dividing by 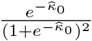.

### S3 OneK1K Genotyping

The genotype IDAT files were parsed by GenomeStudio software and converted to plink files in the genome build of GRCh38. SNPs were quality-controlled by Hardy–Weinberg Equilibrium (HWE) P-value *>* 1 *×* 10^*−*7^, missingness rate *<* 0.05, and minor allele frequency (MAF) *>* 0.01. Individual-wise missingness rate was also controlled by *<* 0.1. The genotypes were further imputed through the Michigan Imputation Server. Imputed SNPs with imputation *r*^2^ *>* 0.5 were retained. Post-imputation QC was also carried out on SNPs by the HWE, missingness, and MAF using the same parameters. After imputation and QCs, the genotype data consist of 1,102 individuals and 4,749,893 SNPs on 22 autosomes and the X chromosome.

### S4 OneK1K Cell Clusters and Trajectories

The single-cell gene expression data were processed by the R package `Seurat` (version 5.2.1). The gene expression count data was log-transformed, then scaled and centered. Top 2,000 differentially-expressed genes were selected for inferring trajectories. Cell clusters were identified through a shared nearest neighbor (SNN) modularity optimization based clustering algorithm, where 20 nearest neighbors for each cell were identified using Euclidean distance. The naïve B cluster and memory B cluster were identified by signature genes *TCL1A, FCER2, IL4R*, and *CD27*. The number ratio of naïve B cells to memory B cells was approximately 2:3, leading to a cell stage pseudotime cutoff at 0.4 in **Figure 4**. Principle component analysis (PCA) was then carried out. Top 50 PCs were used for building the UMAP plot, and 10 neighboring points were used in local approximations of manifold structure. We then used the R package `slingshot` to link the cluster centers and inferred the trajectories. Cell pseudotime is defined as the rank of a cell along the trajectory and then normalized into [0, 1]. Details of `Seurat` and `slingshot` parameters for B cells can be found in Table 4.

**Table 4:**
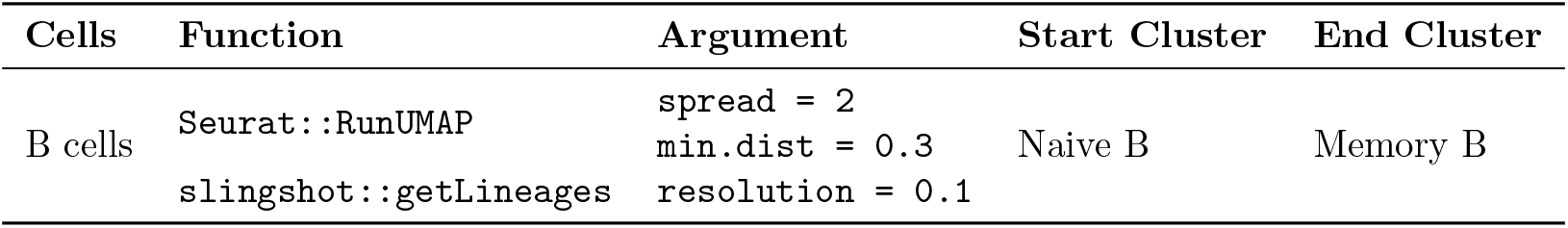
Seurat and slingshot Parameters.

### S5 Pseudotime-dependent TWAS and Pseudobulk TWAS

After cell trajectories and pseudotime were inferred, we removed the subjects with a genotyping missing rate larger than 0.05 or with fewer than 10 single cells per trajectory, resulting in the subject sizes of 965 for B-ALL analysis. All analyzed genes passed the filtering that they had non-zero expression in at least 1% of cells.

For the B-ALL, we screened a list of candidate genes from previous literature (Chokkalingam et al., 2013; Xu et al., 2013; Migliorini et al., 2013; Ellinghaus et al., 2012). The genes include *ARID5B, BMI1, COMMD3, GATA3, LHPP, PIP4K2A, PTPRJ, C14orf119, CEBPE, SLC7A8, TP63, DDC, FIGNL1, IKZF1, INTS10, CDKN2A*, and *CDKN2B*. The genes of interest were analyzed marginally as in Algorithm 1. The overall type I error is controlled by Bonferroni correction. Specifically, for the cell aggregation, we applied the Jenks natural breaks optimization with cluster number *L* = 10. In the gene expression imputation model, we included the common cis-SNPs of the gene, that is, SNPs with minor allele frequency larger than 0.05 and within 1 Mb upstream to the gene start and 1 Mb downstream to the gene end. Adjusted covariates included the sex and top 10 genetic PCs. After gene expression imputation models were computed, we filtered out non-heritable genes with cross-validated heritability smaller than 0.01. The Stage 2 regression was then carried out using SMILES individual-level GWAS data.

Pseudobulk TWAS used the same SNPs and covariates as pt-TWAS, and the outcome was the CPM (counts per million) reads after aggregating the cells per individual. The pseudobulk gene imputation model was fitted using a lasso penalty, carried out by the R package `glmnet`. Similarly, the non-heritable genes with cross-validated heritability smaller than 0.01 were filtered out.

To construct the simultaneous confidence bands, we ran 2,000 parametric bootstrap iterations. We tested a sequence of values for 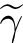 corresponding to the 0.975, 0.980, 0.985, 0.990, 0.995, 0.9995, 0.99995 percentiles of the standard normal distribution, selecting the first value that achieved a simultaneous coverage probability greater than 0.95.

The step spline basis used for significant genes are defined as

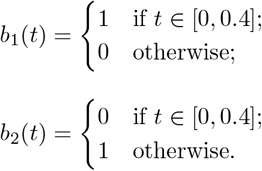

**Figure S1:**
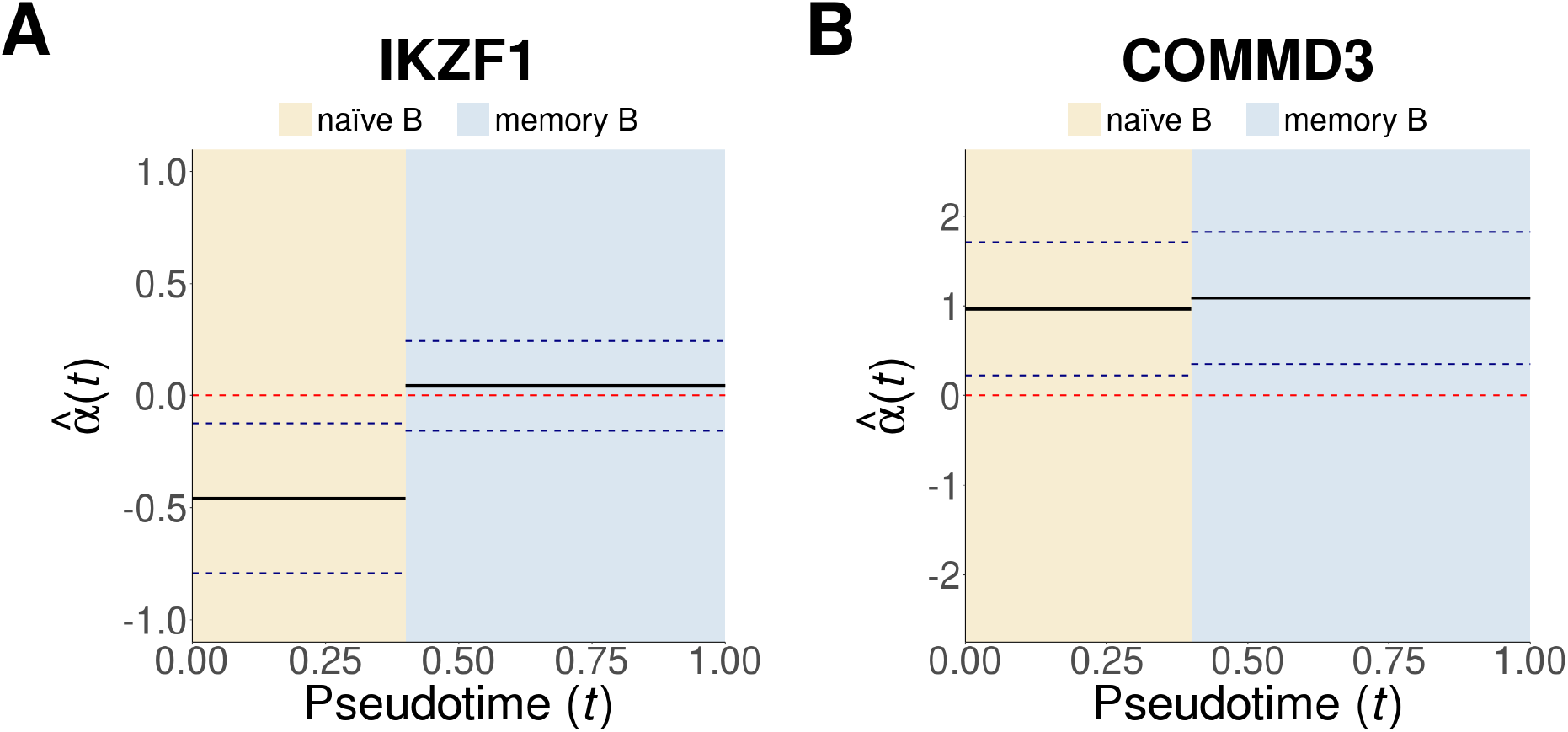
Sensitivity analysis on *IKZF1* (left) and *COMMD3* (right) confidence bands on B-ALL. The coefficient curve *α*(*t*) is estimated under a step function basis. Solid black line,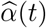; dashed blue line, simultaneous confidence band; dashed red line, horizontal line at 0; beige color, naïve B cell stage; light blue color, memory B cell stage.

## Notes

### Competing Interest Statement

The authors have declared no competing interest.

### Funding Statement

R. Cao gratefully acknowledges support from the Doctoral Dissertation Fellowship by the University of Minnesota. T. Yang is supported by the St.∼Baldrick Career Award (1463960) and NIH grant R01AG074858. C. Li is supported in part by NSF grant DMS-2515789.

### Author Declarations

Ethics committee/IRB of University of Minnesota gave ethnical approval for this work (STUDY00002782).

